# Effectiveness of Face Masks in Blocking the Transmission of SARS-CoV-2: a Preliminary Evaluation of Masks Used by SARS-CoV-2-Infected Individuals

**DOI:** 10.1101/2021.06.20.21259167

**Authors:** Vinicius M. Mello, Cristiane M. Eller, Andreza L. Salvio, Felipe F. Nascimento, Camila M. Figueiredo, Emanuelle S. R. F. Silva, Paulo S. F. Sousa, Pamela F. Costa, Anne A. P. Paiva, Maria A. M. M. Mares-Guias, Elba R. S. Lemos, Marco A. P. Horta

## Abstract

In 2019, a novel severe acute respiratory syndrome coronavirus 2 (SARS-CoV-2), which is transmitted via airborne route, caused a new pandemic namely, “coronavirus disease 2019” (COVID-19). Although it is still debated whether the use of masks can prevent the transmission of SARS-CoV-2, no study has evaluated the virus-blocking efficacy of masks used by patients. We aimed to evaluate this efficacy of masks used by SARS-CoV-2-infected individuals. Data, masks used, and nasopharyngeal swab samples were obtained from these patients. Forty-five paired samples of nasopharyngeal swabs and masks were obtained and processed; the majority of masks were woven. Viral RNAs were amplified using quantitative reverse-transcription polymerase chain reaction and detected only on the inner parts of masks. Median cycle threshold (Ct) values of swabs and masks were 28.41 and 37.95, respectively. Statistically, there was a difference of approximately 10 Ct values between swabs and masks and no significant difference in Ct values among different types of masks. There were statistically significant differences in Ct values between men and women and symptomatic and asymptomatic patients. Our findings suggest the blocking of the transmission of viruses by different types of masks and reinforce the use of masks by both infected and non-infected individuals.

## 1. Introduction

In 2019, a new respiratory coronavirus namely, severe acute respiratory syndrome coronavirus 2 (SARS-CoV-2), which is transmitted via the airborne route, primarily through respiratory droplets and aerosols, caused the new global pandemic associated with a respiratory syndrome namely, the “coronavirus disease 2019” (COVID-19) that has resulted in millions of deaths [1,2] Some studies suggest that the use of a mask can possibly prevent the transmission of several respiratory viruses, such as influenza and rhinovirus, in addition to the new coronavirus [3–5]. Although there has been much discussion regarding whether masks should be used to prevent viral transmission during the initial period of the COVID-19 pandemic, now there is a global understanding of the importance of using masks for preventing SARS-CoV-2 infection. It has been reported that a mask not only protects the person who is wearing it but also reduces the likelihood of transmission of the disease from the person wearing the mask to another person [6].

Current epidemiological data indicate that wearing a mask can reduce the emission of SARS-CoV-2 particles into the environment [7]. The surgical mask (non-woven mask) had a greater filtration efficiency for viral aerosols; however, the filtration efficiency was inferior to that of an N95 mask [8–11]. With the worsening of the pandemic in some countries, especially the developing ones, countries have suffered from the non-availability of surgical masks [8,12,13]. As a great alternative, homemade fabric masks have become very popular in several affected countries, mainly in Brazil [14–20]. Although fabric masks provide less protection and have low filtering efficiency when compared with surgical masks, they may have some effectiveness in preventing the transmission of SARS-CoV-2 [8–11]. Nevertheless, these homemade masks are produced by small-scale fashion productions and do not have quality certifications from health authorities [14–20].

Despite the World Health Organization recommendations about the use of face masks, it is still controversial whether it reduces the risk of transmission of SARS-CoV-2 [21]. No study has evaluated the presence of retained viruses on the masks, which are made using different materials and used by SARS-CoV-2-infected individuals, and the effectiveness of these masks in preventing viral transmission. Considering the heterogeneity of cloth masks that are sold in Brazil, it is still unclear whether these homemade masks are effective in blocking the transmission of virus. Considering these points, in the present study, we aimed to evaluate the virus-blocking efficacy of masks used by SARS-CoV-2-infected individuals.

The results presented here suggest that the use of masks helps to block viral transmission by SARS-CoV-2-infected individuals and reinforce the importance of using masks as a preventive measure against the viral transmission.

## 2. Materials and Methods

### 2.1. Ethics Statement

The Oswaldo Cruz Institute/IOC/FIOCRUZ Research Ethics Committee approved this study (number: CAAE 37142520.0.0000.5248). All procedures were performed in accordance with the ethical standards of the responsible committees on human experimentation (institutional and national) and the Helsinki Declaration of 1975, as revised in 2008. All patients who were included in the study agreed with their participation in the research by signing the informed consent.

### 2.2. Study Population and Sample Collection

Nasopharyngeal swab samples and masks were collected (between December 2020 to March 2021) from patients who were suspected to be infected by SARS-CoV-2 and attended the Municipal Theatre and Benjamin Constant Institute survey, conducted in the city of Rio de Janeiro, Brazil, according to medical decision and after obtaining permissions from the patients.

Samples were collected as follows: a nasopharyngeal swab was inserted in the nostril until it hit an obstacle (the inferior concha or the back of the nasopharyngeal cavity), rotated, and removed. The test was conducted in two nostrils per patient. After sampling, the nasopharyngeal swab was inserted into a vial containing 3 mL of a viral transport medium (VTM; Xpert nasopharyngeal sample collection kit, Cepheid, Sunnyvale, CA, USA). After the collection of swab samples, the masks used for 2–3 h by the participants were placed inside a clean plastic bag and they were provided clean, new masks for use. Furthermore, data, including the biological sex and age of these patients were collected.

### 2.3. Processing of Masks and Swabs

Nasopharyngeal swab specimen was collected and immediately resuspended in 3 mL of the VTM. For mask samples, immediately after the collection of masks, pieces were cut based on the following reference measures: the right side and left side areas with a width of 2 cm each, obtained after removing side seam using the entire height of the mask; the nose area (N) with a height of 5 cm and width of 5 cm; and the mouth area (M) with a height of 5 cm and width of 8 cm, and subsequently, these pieces were added to the VTM. In cases of samples with double or triple layers of the material, these areas were subdivided into inner part of N, middle part of N, outer part of N, inner part of M, middle part of M, and outside part of M, respecting the sizes of the cut areas previously described **(Figure 1)**.

**Figure 1.**
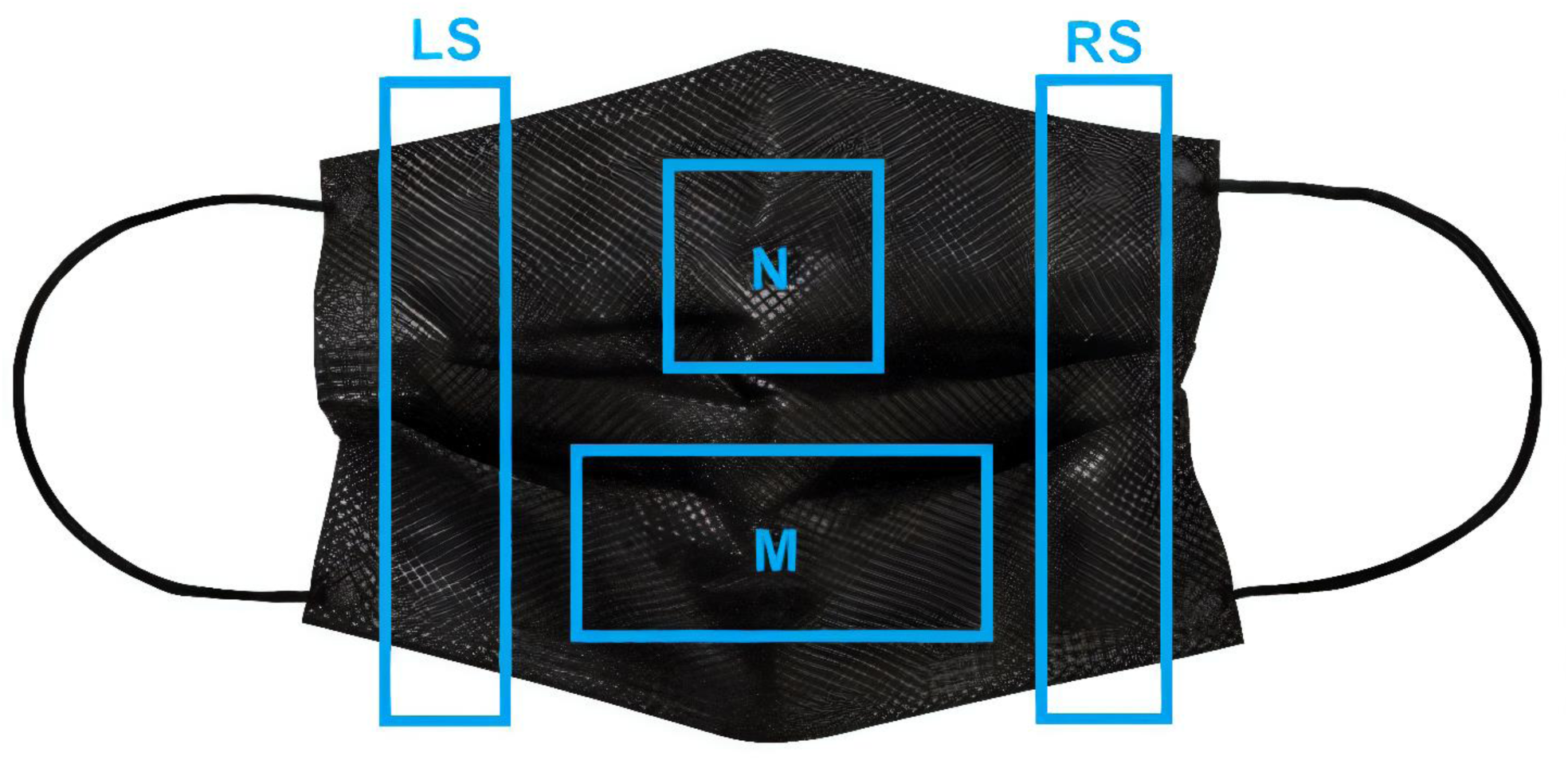
The scheme for cutting a mask. The areas enclosed within blue margins represent the areas cut from the mask. LS = Left Side, RS = Right Side, M = Mouth area, and N = Nose area.

Between resuspension and the processing of each sample (swabs and mask pieces), incubation at 4 °C for a minimum of 30 minutes and a maximum of 12 hours was be performed. Subsequently, the samples were processed through vortex homogenization and transferred from the medium to a previously identified 1.5-mL tube using a Pasteur pipette (2 mL). Then, swabs and masks were discarded, and the final sample in the medium was stored at -80 °C.

### 2.4. Viral Genome Extraction

Nucleic acids from all the samples were extracted and purified using the DNA/RNA 300 kit H96 in the Janus G3 and Janus Chemagic automatic extractor (Perkin-Elmer, Waltham, USA). The Janus 360 system is based on magnetic spheres for extracting viral nucleic acids from 300 uL of the sample. The operation of the equipment and the use of the commercial kit were in accordance with the manufacturer’s instructions.

### 2.5. SARS-CoV-2 Molecular Detection

For SARS-CoV-2 genome amplification, we used a molecular kit for the E region (Bio-Manguinhos, Rio de Janeiro, BR) following the manufacturer’s instructions. The plate setup was automated and performed using Janus G3 (Perkin-Elmer, Waltham, USA). In this method, the quantitative reverse-transcription polymerase chain reaction (qRT-PCR) also allowed the quantification of viral genomic RNA of SARS-CoV-2 with the application of an in-house ssRNA standard curve. The chosen commercial kit helped in detecting the E region of the genome using a FAM probe and the RP human gene using a VIC probe; the latter functions as the internal positive control of the assay. For all assays, positive and negative controls were included in the commercial molecular kit, and they were used in all experiments.

Samples with a cycle threshold (Ct) value lower than 38.0 for E region were considered positive, and negative samples were the ones that presented a Ct value higher than 38.0 or no Ct value at all. For the RP target, a Ct value equal to or lower than 35.0 validated the experiment. The positive control Ct value must be lower than 37.0 to validate the assay.

### 2.6. Statistical Analysis

The results of descriptive statistical analyses are presented using frequency tabulations and percentages. Medians are presented with interquartile range (IQR) values. The Mann–Whitney U test was used to compare the differences in Ct values between the independent groups of masks and swabs. Statistical significance was set at a p-value ≤ *0.05*. All analyses were performed using R software version 4.1.0 (The R Foundation for Statistical Computing, Vienna, Austria).

## 3. Results

Forty-five swab samples with their paired respective masks were collected. The masks were classified as woven masks (30/45; 66.7%) and surgical non-woven masks (15/45, 33.3%). SARS-CoV-2 RNA was detected in all swab samples and masks. The viral RNA was detected only on the inner part (the part that was in contact with the face) of the masks. None of the masks was positive for the RNA on the outer part (the part that was in contact with the external environment). The median Ct values of the swab and mask samples were 28.41 (IQR, 21.55–31.74) and 37.95 (IQR, 33.50 – 40.00), respectively. The descriptive information can be seen in **Table 1**.

**Table 1.**
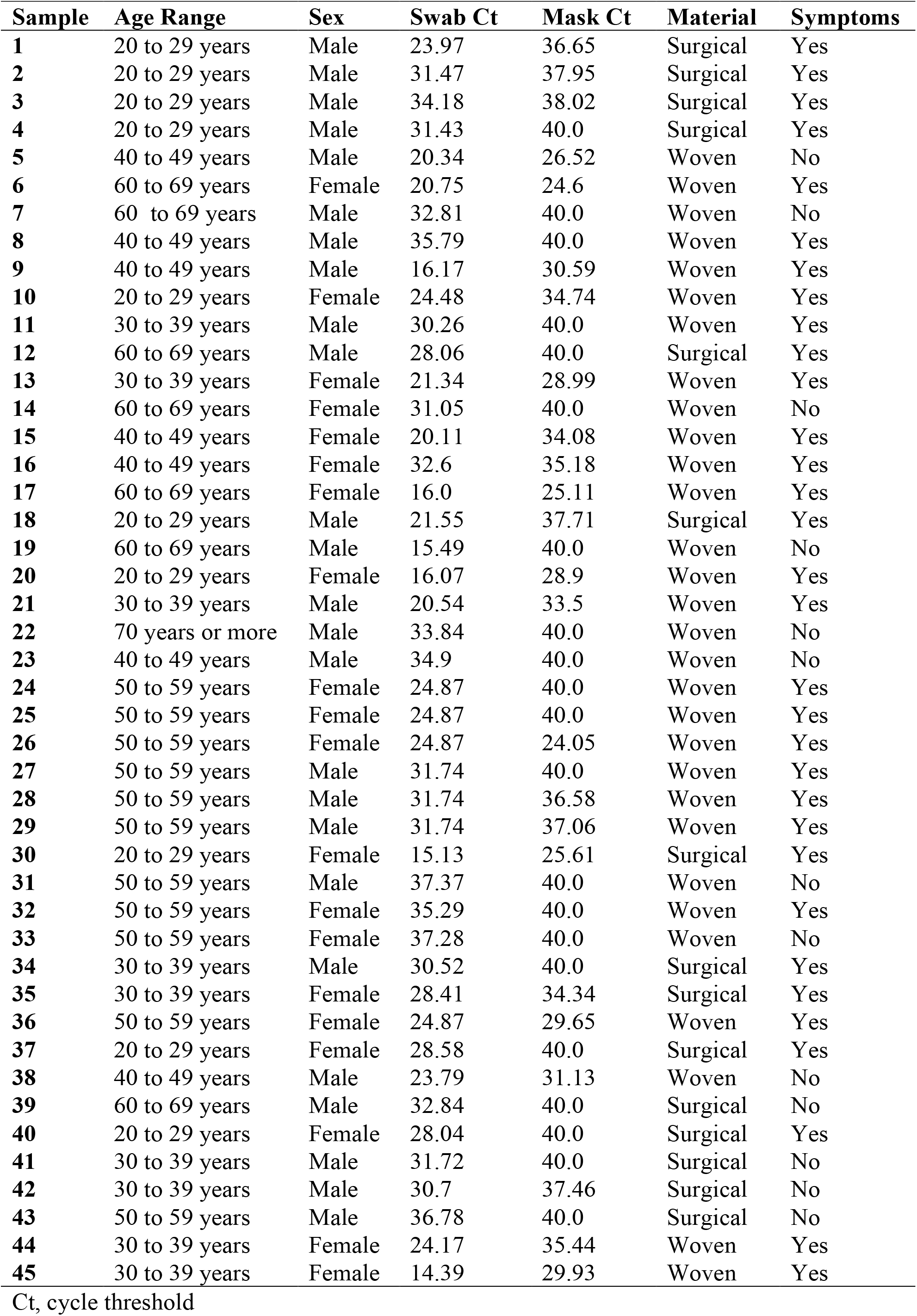
Clinical features of the patients infected by SARS-CoV-2 in the present study

Our analysis showed a reduction of approximately 10 Ct values (≅3 logs or 1000 RNA copies/mL) between swab and mask samples. Statistical analysis considering the adjusted linear equation showed a relationship in reduction in Ct values (Y = 0.54X + 21.42), with a positive and significant correlation (rho = 0.64, p<0.001) (Figure 2).

**Figure 2.**
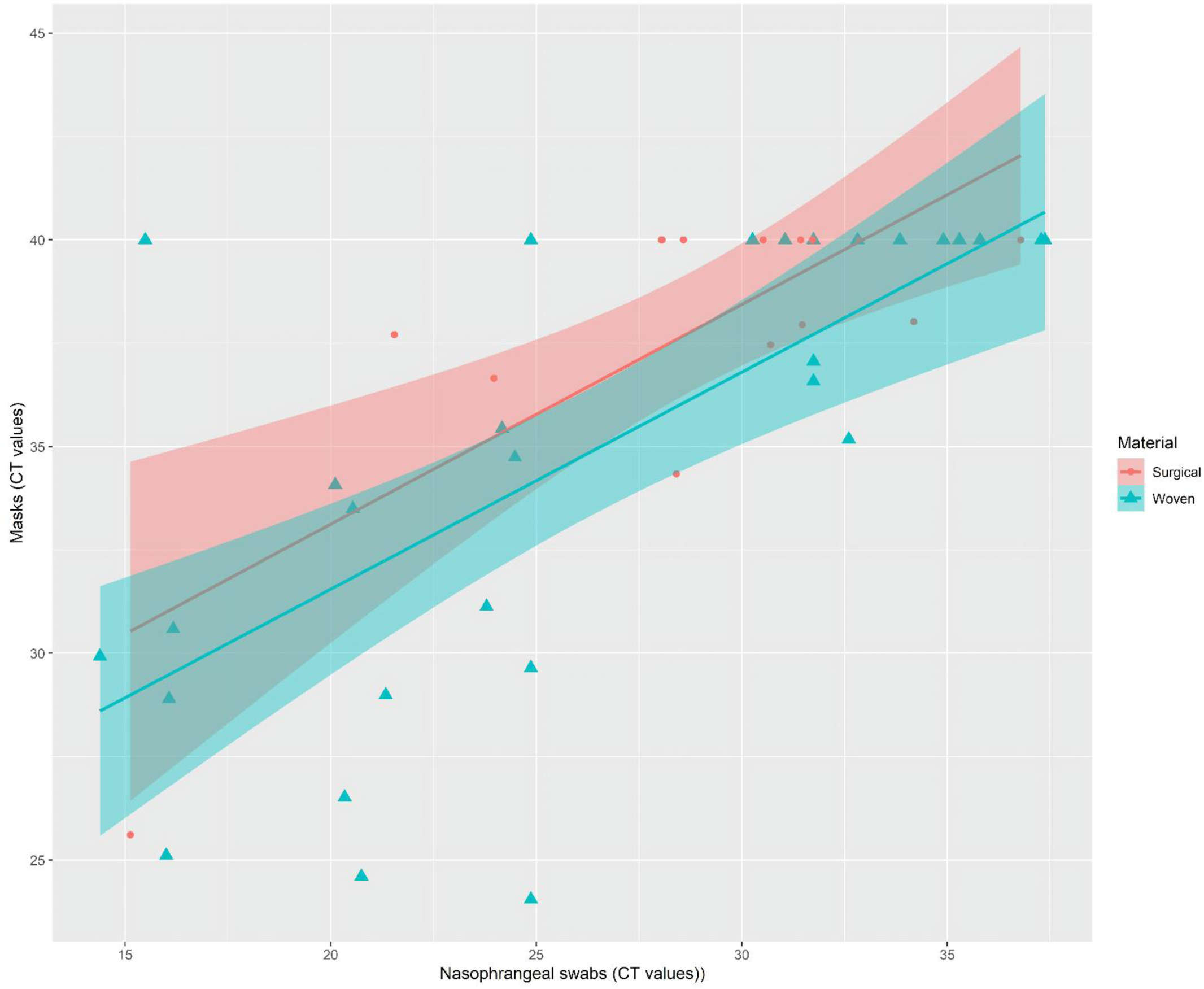
Relationship between severe acute respiratory syndrome coronavirus 2 viral loads (Ct values) of nasopharyngeal swabs and masks used by infected patients. Ct value, cycle threshold value.

The analysis did not identify a statistically significant difference in median Ct values between surgical and cloth masks (U = 266.5, *p = 0.12*). The same result was obtained when comparing Ct values of nasopharyngeal swabs (U = 187, *p = 0.56*). Women and men presented significant differences in median Ct values of masks (U = 150, *p = 0.01*) and swabs (U = 151, *p = 0.02*). We observed a significant difference in Ct values between masks used by patients with symptoms of COVID-19 and those used by patients with no clinical manifestations of COVID-19 (U = 265, *p = 0.004*). Further results of statistical analysis can be found in **Table 2**.

**Table 2.** Median cycle threshold values of masks and nasopharyngeal swabs from 45 patients infected by SARS-CoV-2

|  | N (%) | Masks Ct | IQR | *P-value* | Swab Ct | IQR | *P-value* |
| --- | --- | --- | --- | --- | --- | --- | --- |
| **Sex** |  |  |  | *0.01* |  |  | *0.02* |
| *Female* | 20 (44.4) | 34.54 | (28.69–40.00) |  | 24.87 | (20.5–28.45) |  |
| *Male* | 25 (55.6) | 40.00 | (28.69–40.00) |  | 31.47 | (23.97–32.84) |  |
| **Mask type** |  |  |  | *0.11* |  |  | *0.40* |
| *Surgical* | 15 (33.3) | 40.00 | (37.58–40.00) |  | 30.52 | (28.05–31.59) |  |
| *Woven* | 30 (66.7) | 36.01 | (30.09–40.00) |  | 24.87 | (20.59–32.38) |  |
| **Symptoms** |  |  |  | *0.004* |  |  | *0.004* |
| *Yes* | 32 (71.1) | 36.58 | (30.26–40.00) |  | 24.87 | (21.04–31.45) |  |
| *No* | 13 (28.9) | 40.00 | (40.00–40.00) |  | 32.82 | (30.96–35.37) |  |

IQR = Interquartile range; Ct, cycle threshold value.

## 4. Discussion

After 1 year of the COVID-19 pandemic, the Americas have become the epicenter of COVID-19 cases and deaths; especially in Brazil, there has been an increase in the average number of deaths [1]. This may be associated with late interventions against the pandemic and adherence to scientific negationism, for example, not wearing protective masks, among other factors [22,23].

The results in this study, reinforce the evidence that in general, wearing masks can be beneficial to the community and that this beneficial effect is derived from the ability of masks to block the exhalation and inhalation of infectious viruses, regardless of the type of mask used, as shown in a review by Brooks and Butler (2021) [24]

Data from different studies conducted in several countries have shown that the use of masks together with social distancing can reduce the transmission of SARS-CoV-2 and the number of cases of SARS-CoV-2 infection [11,25–28]. A study performed by Ma et al. (2020), which used an automated system that mimicked human breathing, showed that the virus-blocking rates of surgical and homemade masks were approximately 97% and 95%, respectively [29]. Another study performed by Morais et al. (2021), which used a similar methodology for evaluating different mask types, demonstrated similar results, where surgical masks had a filtration rate of 89% and homemade masks had filtration rates ranging from 40% to 83%, depending on the type of the fabric [30]. In the present study, we observed a reduction by approximately 10 Ct values (≅3 logs [≅x1000 RNA copies/mL]) for masks compared with the paired swabs collected from the same individual. These findings corroborate with data from the previous studies [29,30] that indicate a possible blocking of viral transmission by masks worn by infected people; these results may shed light on the effectiveness of masks in blocking SARS-CoV-2.

Another result that reinforced the hypothesis of blocking of viral transmission by masks was that only inner parts (the parts in contact with the face) of the masks were positive for viral RNA. Furthermore, the reduction in Ct values showed a significant statistical association (rho = 0.64, p<0.05), showing that the virus-blocking rate is possibly relevant in preventing the transmission of the virus from infected people to other individuals, being bowed with results found in the literature [24].

It is important to highlight that a reduction in Ct values was observed in the different types of masks (non-woven and woven masks) analyzed and we compared swab samples and masks, which were collected simultaneously. Additionally, there was no statistically significant difference in the decrease in Ct values among different types of masks. These results reveal that different types of masks may be used to reduce the transmission of viruses to the environment and prevent infection in susceptible individuals. A similar result was obtained in a study performed by Zangmeister et al. (2020) that evaluated the effectiveness of the materials of cloth masks, which were used to reduce the transmission of SARS-CoV-2, in the filtration of nanoscale aerosols and showed that no cloth mask performed similar to an N95 mask. However, woven, and non-woven cloth masks may be used to reduce the transmission of SARS-CoV-2 and to filter viral particles [31].

In this context, in a country like Brazil, where it is impossible to totally adopt measures of social distancing, mainly in socially vulnerable populations in peripheral areas and slums, the use of masks seems essential. Moreover, the use of masks could be beneficial to those individuals who still need to use public transport, such as buses, trains, and/or subways, which are often crowded [21,32–34]. The use of masks, especially woven ones, is extremely relevant as an additional protective measure for reducing the increasing number of cases and deaths due to COVID-19 in Brazil [12].

Statistically significant results (p = 0.02 and p = 0.01, respectively) were obtained on comparing Ct values between swabs and masks from men and women. This may be directly associated with the sex—a hypothesis to be considered is a greater release of viral particles by males. Some studies have shown that males have a significantly high risk of severe disease, mainly due to differences in inflammatory responses to viral infections, and genetic and hormonal regulation [35–37]. However, more studies are needed to understand the underlying biological phenomena.

Some studies suggest that the viral load found in asymptomatic patients is similar to that found in symptomatic patients [38–40]. However, we identified lower Ct values in symptomatic patients than those in asymptomatic patients, and this difference was statistically significant (p = 0.004), indicating an elevated viral load mainly in swab samples.

This was a preliminary study and has some limitations. The sample size was relatively small, and it did not evaluate the filtering efficiency of the masks as performed in some other studies [28,29]. Furthermore, this study only evaluated masks from SARS CoV 2 infected individuals with a positive qRT-PCR. Further studies are needed to evaluate the masks of uninfected individuals who have direct contact with infected individuals. Further studies, including a larger number of masks, are also needed to analyze the viability of the virus detected in infected masks through cell culture.

However, our results provided real-life evidence regarding blocking of viral transmission by masks used by individuals infected by SARS-CoV-2. Furthermore, the results also reinforce the suggestion to use a mask by everyone, regardless of whether the individual is infected or not. This is important since there are asymptomatic cases of infection and evidence of transmission of the virus even before the appearance of the first symptoms, as reported by some studies [38,41].

## 5. Conclusions

The study results shed light on the importance of using masks by individuals infected with SARS-CoV-2 and shows that different types of masks can help block viral transmission. Moreover, our findings also reinforce the importance of using masks as a preventive measure against the viral transmission, regardless of the type of mask used, in addition to social distancing and personal hygiene measures.

## Data Availability

I agree to provide the data if necessary and requested

## Author Contributions

V.M.M., A.L.S., M.A.P.H.: Writing—original draft preparation; Concep-tualization. V.M.M., A.L.S., C.M.E., M.A.P.H.: writing—review and editing. C.M.E., A.L.S., C.M.F: sample processing, data curation, methodology. C.M.E., A.L.S., C.M.F, P.S.F.S.: sample collection. F.F.N.: Mask cut design and development, sample processing, methodology, sample collection. E.S. R. F.S., P.S.F.S., P. F.C., A.A.P.P., M.A.M.M.M-G.: methodology. E.R.S.L.: review, editing, conceptualization, investigation, project administration, funding acquisition. M.A.P.H.: data processing, funding acquisition, project administration. All authors have read and agreed to the published version of the manuscript.

## Institutional Review Board Statement

The study was conducted according to the guidelines of the Declaration of Helsinki and approved by Ethics Committee of the Oswaldo Cruz Insti-tute/IOC/FIOCRUZ (protocol code: CAAE 37142520.0.0000.5248 approved on September 30, 2020).

## Informed Consent Statement

All patients in the study were aware of and according to their par-ticipation in the research, after signing the informed consent.

## Acknowledgments

We are also grateful to the Rio de Janeiro Municipal Theatre team and Ben-jamin Constant Institute team for our collaboration. We especially thank all people who made themselves available to participate in this study through the swab and mask collections.

## References

1. World Health Organization. WHO Coronavirus (COVID-19) Dashboard 2021. https://covid19.who.int. Accessed March 31, 2020.

2. Harrison AG, Lin T, Wang P. Mechanisms of SARS-CoV-2 Transmission and Pathogenesis. Trends Immunol. 2020;41(12):1100–15. doi: 10.1016/j.it.2020.10.004

3. Leung NHL, Chu DKW, Shiu EYC, et al. Respiratory virus shedding in exhaled breath and efficacy of face masks. Nat Med. 2020; 26(5):676–680. DOI: 10.1038/s41591-020-0843-2.

4. Milton DK, Fabian MP, Cowling BJ, et al. Influenza virus aerosols in human exhaled breath: particle size, culturability, and effect of surgical masks. PLoS Pathog. 2013;9(3):e1003205. doi: 10.1371/journal.ppat.1003205.

5. Prather KA, Wang CC, Schooley RT. Reducing transmission of SARS-CoV-2. Science. 2020;368(6498):1422–4. doi: 10.1126/science.abc6197.

6. Swain ID. Why the mask? The effectiveness of face masks in preventing the spread of respiratory infections such as COVID-19 - a home testing protocol. J Med Eng Technol. 2020;44(6):334–7. doi: 10.1080/03091902.2020.1797198.

7. Střížová Z, Bartůňková J, Smrž D. Can wearing face masks in public affect transmission route and viral load in COVID-19? Cent Eur J Public Health. 2020;28(2):161–2. doi: 10.21101/cejph.a6290.

8. World Health Organization. Mask use in the context of COVID-19. 2020. https://apps.who.int/iris/handle/10665/337199. Accessed April 5, 2020.

9. Guan L, Zhou L, Zhang J, et al. More awareness is needed for severe acute respiratory syndrome coronavirus 2019 transmission through exhaled air during non-invasive respiratory support: experience from China. Eur Respir J. 2020;55(3). doi: 10.1183/13993003.00352-2020.

10. Remuzzi A, Remuzzi G. COVID-19 and Italy: what next? Lancet. 2020;395(10231):1225–8. doi: 10.1016/S0140-6736(20)30627-9.

11. Choi S, Ki M. Estimating the reproductive number and the outbreak size of COVID-19 in Korea. Epidemiol Health. 2020;42:e2020011. doi: 10.4178/epih.e2020011.

12. Ortelan N, Ferreira AJF, Leite L, et al. Cloth masks in public places: an essential intervention to prevent COVID-19 in Brazil. Cien Saude Colet. 2021;26(2):669–92. doi: 10.1590/1413-81232021262.36702020.

13. Ji D, Fan L, Li X, Ramakrishna S. Addressing the worldwide shortages of face masks. BMC Mater. 2020;2(1):9. doi: 10.1186/s42833-020-00015-w.

14. Zhou SS, Lukula S, Chiossone C, et al. Assessment of a respiratory face mask for capturing air pollutants and pathogens including human influenza and rhinoviruses. J Thorac Dis. 2018;10(3):2059–69. doi: 10.21037/jtd.2018.03.103.

15. Offeddu V, Yung CF, Low MSF, Tam CC. Effectiveness of Masks and Respirators Against Respiratory Infections in Healthcare Workers: A Systematic Review and Meta-Analysis. Clin Infect Dis. 2017;65(11):1934–42. doi: 10.1093/cid/cix681.

16. Chua MH, Cheng W, Goh SS, et al. Face Masks in the New COVID-19 Normal: Materials, Testing, and Perspectives. Re-search (Wash D C). 2020;2020:7286735. doi: 10.34133/2020/7286735.

17. Santos M, Torres D, Cardoso PC, et al. Are cloth masks a substitute to medical masks in reducing transmission and contamination? A systematic review. Braz Oral Res. 2020;34:e123. doi: 10.1590/1807-3107bor-2020.vol34.0123.

18. Silva Acoe, Almeida AM, Freire MEM, et al. Cloth masks as respiratory protections in the COVID-19 pandemic period: evidence gaps. Rev Bras Enferm. 2020;73(Suppl 2):e20200239. doi: 10.1590/0034-7167-2020-0239.

19. Sun P, Lu X, Xu C, et al. Understanding of COVID-19 based on current evidence. J Med Virol. 2020;92(6):548–51. doi: 10.1002/jmv.25722.

20. Cowling BJ, Chan KH, Fang VJ, et al. Facemasks and hand hygiene to prevent influenza transmission in households: a cluster randomized trial. Ann Intern Med. 2009;151(7):437–46. doi: 10.7326/0003-4819-151-7-200910060-00142.

21. Cheng VC, Wong SC, Chuang VW, et al. The role of community-wide wearing of face mask for control of coronavirus disease 2019 (COVID-19) epidemic due to SARS-CoV-2. J Infect. 2020;81(1):107–14. doi: 10.1016/j.jinf.2020.04.024.

22. Kibuuka BGL. Complicity and Synergy Between Bolsonaro and Brazilian Evangelicals in COVID-19 Times: Adherence to Scientific Negationism for Political-Religious Reasons. International Journal of Latin American Religions. 2020;4(2):288–317. DOI: 10.1007/s41603-020-00124-0.

23. Massarani, L, Neves, LFF. Communicating the” race” for the COVID-19 vaccine: an exploratory study in newspapers in the United States, the United Kingdom and Brazil. 2021; 4:41. DOI: 10.3389/fcomm.2021.643895.

24. Brooks JT, Butler JC. Effectiveness of Mask Wearing to Control Community Spread of SARS-CoV-2. JAMA. 2021;325(10):998–9. doi: 10.1001/jama.2021.1505.

25. Centers for Disease Control and Prevention. CDC. 2021. Science Brief: Community Use of Cloth Masks to Control the Spread of SARS-CoV-2. https://www.cdc.gov/coronavirus/2019-ncov/science/science-briefs/masking-science-sars-cov2.html. Accessed April 5, 2020.

26. Clapham HE, Cook AR. Face masks help control transmission of COVID-19. Lancet Digit Health. 2021;3(3):e136–e7. doi: 10.1016/S2589-7500(21)00003-0.

27. Mitze T, Kosfeld R, Rode J, Wälde K. Face masks considerably reduce COVID-19 cases in Germany. Proc Natl Acad Sci U S A. 2020;117(51):32293–301. doi: 10.1073/pnas.2015954117.

28. Chu DK, Akl EA, Duda S, et al. Physical distancing, face masks, and eye protection to prevent person-to-person trans-mission of SARS-CoV-2 and COVID-19: a systematic review and meta-analysis. Lancet. 2020;395(10242):1973–87. doi: 10.1016/S0140-6736(20)31142-9.

29. Ma QX, Shan H, Zhang HL, et al. Potential utilities of mask-wearing and instant hand hygiene for fighting SARS-CoV-2. J Med Virol. 2020;92(9):1567–71. doi: 10.1002/jmv.25805.

30. Morais FG, Sakano VK, de Lima LN, et al. Filtration efficiency of a large set of COVID-19 face masks commonly used in Brazil. Aerosol Sci Techno. just-accepted, 2021:1–15.

31. Zangmeister CD, Radney JG, Vicenzi EP, Weaver JL. Filtration Efficiencies of Nanoscale Aerosol by Cloth Mask Materials Used to Slow the Spread of SARS-CoV-2. ACS Nano. 2020;14(7):9188–9200. doi: 10.1080/02786826.2021.1915466.

32. Fernandes LAC, Silva CAF, Dameda C, Bicalho PPG. Covid-19 and the Brazilian Reality: The Role of Favelas in Combating the Pandemic. Front. Sociol. 2020;5:611990. doi: 10.3389/fsoc.2020.611990.

33. United Nations. Brazil’s favelas organize to fight Covid-19 2021. https://www.un.org/en/coronavirus/brazil%E2%80%99s-favelas-organize-fight-covid-19. Accessed April 9, 2020

34. Pereira RJ, Nascimento Gnld, Gratão LHA, Pimenta RS. The risk of COVID-19 transmission in favelas and slums in Brazil. Public Health. 2020;183:42–3. doi: 10.1016/j.puhe.2020.04.042.

35. Griffith DM, Sharma G, Holliday CS, et al. Men and COVID-19: A Biopsychosocial Approach to Understanding Sex Dif-ferences in Mortality and Recommendations for Practice and Policy Interventions. Prev Chronic Dis. 2020;17:E63. doi: 10.5888/pcd17.200247.

36. Peckham H, de Gruijter NM, Raine C, et al. Male sex identified by global COVID-19 meta-analysis as a risk factor for death and ITU admission. Nat Commun. 2020;11(1):6317. doi: 10.1038/s41467-020-19741-6.

37. Bienvenu LA, Noonan J, Wang X, Peter K. Higher mortality of COVID-19 in males: sex differences in immune response and cardiovascular comorbidities. Cardiovasc Res. 2020;116(14):2197–2206. doi: 10.1093/cvr/cvaa284.

38. Huff HV, Singh A. Asymptomatic Transmission During the Coronavirus Disease 2019 Pandemic and Implications for Public Health Strategies. Clin Infect Dis. 2020;71(10):2752–2756. doi: 10.1093/cid/ciaa654.

39. Zou L, Ruan F, Huang M, et al. SARS-CoV-2 Viral Load in Upper Respiratory Specimens of Infected Patients. N Engl J Med. 2020 Mar 19;382(12):1177–1179. DOI: 10.1056/NEJMc2001737.

40. Dhama K, Khan S, Tiwari R, et al. Coronavirus Disease 2019-COVID-19. Clin Microbiol Rev. 2020; 33 (4): e00028–20. DOI: 10.1128/CMR.00028-20.

41. Buitrago-Garcia D, Egli-Gany D, Counotte MJ, et al. Occurrence and transmission potential of asymptomatic and pre-symptomatic SARS-CoV-2 infections: A living systematic review and meta-analysis. PLoS Med. 2020;17(9): e1003346. Published 2020 Sep 22. DOI: 10.1371/journal.pmed.1003346.

